# Higher Ventilation Rate is Associated with Increased Return of Spontaneous Circulation in In-Hospital Cardiac Arrest Patients with Advanced Airways

**DOI:** 10.1101/2025.04.17.25326045

**Authors:** Ian S. Jaffe, Yulan Ren, Linh Tran, Eugene Yuriditsky, Anelly M. Gonzales, Jignesh K. Patel, Samia Shahnawaz, James Horowitz, Ben Bloom, Deepak Pradhan, Erik Kulstad, Heather Jarman, Nam Tong, Matthew Thomas, Louisa Chan, Valerie Page, Charles Deakin, Gavin D. Perkins, Chang Yu, Sam Parnia

## Abstract

**Background:** Current cardiopulmonary resuscitation (CPR) guidelines recommend a ventilation rate of 10 breaths/min in adult cardiac arrest patients with an advanced airway in place; however, this guideline is based almost exclusively on animal model studies. The objective of this study was to examine the association between mean ventilation rates during in-hospital cardiac arrest with return of spontaneous circulation (ROSC).

**Methods:** This was a secondary analysis of a cohort undergoing CPR for in-hospital cardiac arrest with an advanced airway in place had that ventilation rate and end-tidal CO_2_ (ETCO_2_) continuously recorded. Subjects were enrolled at 25 tertiary care centers in the United States and United Kingdom. A subset of subjects also had intra-cardiac arrest arterial blood gases collected as part of routine clinical care.

**Results:** Ventilation rate and ETCO_2_ measurements were collected for 222 subjects and arterial blood gas data for 127 subjects. We observed 84.7% of subjects were ventilated > 10 breaths/min. When dichotomized to subjects receiving close to current guidelines (6-12 breaths/min) and those with higher ventilation rates >12 breaths/min, subjects with higher ventilation rates had greater rate of ROSC (45% vs. 24% p = 0.009). Ventilation rate had a significant impact on ROSC even after adjusting for age, sex, shockable initial rhythm, pre-cardiac arrest severity of critical illness, and mechanical chest compression device usage, with an adjusted odds ratio of 1.15 per two breaths/min increase (95% CI 1.04-1.28; p = 0.006). Regression analysis suggested increasing ventilation rates may start to have a negative association with ROSC when beyond 26 breaths/min. Patients ventilated > 12 breaths/min demonstrated increased mean ETCO_2_ levels (median 25 mm Hg vs. 17 mm Hg for those with mean ventilation rates within 6-12 breaths/min; p < 0.001). PaO_2_ and PaCO_2_ levels did not differ between ventilation groups, suggesting that the observed effects may relate to hemodynamic rather than ventilatory effects.

**Conclusions:** Ventilation in excess of guideline-recommended rates is common. Higher ventilation rates above 6-12 breaths and below 26 breaths/min were associated with increased odds of ROSC. This may be due to improved hemodynamics. Guidelines on ventilation during CPR may require further examination.

## Introduction

There are approximately 650,000 resuscitation attempts for cardiac arrest in the United States annually, with in-hospital cardiac representing approximately 290,000 resuscitation attempts.^1–3^ Survival rates from in-hospital cardiac arrest are approximately 10-20%, with wide variability depending on the centers studied and quality of CPR delivered.^2,4^ High-quality cardiopulmonary resuscitation (CPR) is essential for survival from cardiac arrest,^5^ although CPR quality is inconsistent, both in and out of hospitals.^2,6–8^ Though recent studies have focused on the use of compression-only CPR in untrained or minimally trained bystanders, advanced CPR has always included mechanical ventilations.^9^ While chest compressions have been studied in many trials, the precise dynamic of ventilation in CPR remains poorly understood.^10–14^ This is especially true in the context of an advanced airway, in which ventilations and compressions are delivered continuously.^15^ In short, ventilations are an important component of CPR when well-trained clinicians are present, but the optimal ventilation rate and volume has not been established. This stems from a paucity of evidence on the role of ventilations in CPR with an advanced airway.

The 2020 American Heart Association Guideline for Cardiopulmonary Resuscitation and Emergency Cardiovascular Care recommend a ventilation rate of 10 breaths/min in adult cardiac arrest patients with an advanced airway in place.^16^ This guideline is largely based on a 2017 systematic review that included a single human observational study (n = 67) and 10 animal studies, which found the recommendation of 10 breaths/min in intubated cardiac arrest patients was based on low-quality evidence.^13^

Ventilation in excess of this recommendation has been observed in numerous observational studies.^17–21^ Animal studies have indicated that hyperventilation during CPR may decrease survival by countering negative intrathoracic pressures generated during chest decompression, leading to decreased venous return and cardiac output, increased pulmonary vascular resistance, and reduced coronary perfusion pressure.^21–24^ At the same time, studies have shown that slow ventilation rates during CPR are associated with hypoxia and hypercapnia, leading to greater metabolic derangements.^25,26^

Recent clinical evidence remains mixed. A 2019 study of adult in-hospital cardiac arrest patients (n = 337) found no significant difference in survival with good neurological outcomes when dichotomizing patients receiving <10 and >10 breaths/min.^27^ A 2017 study of pediatric patients with in-hospital cardiac arrest (n = 52) demonstrated that children resuscitation with a ventilation rate greater than 25 breaths/min were more likely to survive to hospital discharge than patients with a lower ventilation rate (odds ratio [OR]: 4.73; p = 0.029).^28^ This observational study led to the 2020 AHA guideline for pediatric resuscitation changing to recommend an increased ventilation rate of 20-30 breaths/min for pediatric cardiac arrest patients with an advanced airway.^28,29^ Recent evidence in out-of-hospital cardiac arrests has also suggests ventilation at 12-16 breaths/min may be associated with increased odds of favorable neurological survival (OR: 1.36,95% CI 1.01–1.84) compared to ventilation at 6-12 breaths/min, but ventilation >16 breaths/min had no association with increased survival.^30^

In this article we present the results of an observational study of in-hospital cardiac arrests in which continuous ventilatory and ETCO_2_ monitoring were used. Our hypothesis was that a ventilation rate greater than 6-12 breaths/min in patients undergoing CPR with an advanced airway would be associated with higher rates of return of spontaneous circulation (ROSC).

## Methods

### Setting and Design

This study is a cohort analysis of Awareness During Resuscitation II (AWARE II), a prospective multisite observational study of consciousness in cardiac arrest conducted at 25 sites in the United States and United Kingdom.^31^ AWARE II included collection of cardiac arrest data and markers of resuscitation quality using end tidal carbon dioxide (ETCO_2_), regional cerebral oxygenation (rSO2), and electroencephalography (EEG). AWARE II began in 2013 and concluded in March 2020. The cohort of interest for this study was patients enrolled in AWARE II who had continuous end-tidal CO2 (ETCO_2_) and ventilation rate data collected during their resuscitation. This cohort was selected to represent patients who have reached the metabolic phase of cardiac arrest (lasting > 10 minutes); of the three phases of cardiac arrest (electrical, circulatory, and metabolic) patients who reach the metabolic phase have the worst outcomes.^32^

The inclusion criteria for AWARE-II were: Age ≥18 years with in-hospital cardiac arrest with CPR lasting ≥5 min (although most patients enrolled had >10 min of CPR due to the time required to place the monitoring devices used in AWARE-II). Exclusion criteria were: age <18 years, out-of-hospital cardiac arrest events, and repeat in-hospital cardiac arrest events within 14 days of a prior cardiac arrest. The research protocol received ethics committee approval (NYUIRB-s17-00142) across all study sites prior to enrolling the first participant. Patients received CPR in accordance with advanced cardiac life support (ACLS) recommendations active at the time of enrollment (2010 and 2015).^33,34^ Written informed consent was obtained from all survivors; a waiver of consent was approved to use data for non-survivors. Those who survived the initial cardiac arrest were followed up until hospital discharge or death. Dedicated, trained research staff at participating sites attended all in-hospital cardiac arrest events in real-time.

### Study Definitions and Outcome Measures

In this study, in-hospital cardiac arrest is defined as absence of heartbeat and respirations requiring CPR that commenced anywhere in the hospital. Sustained ROSC is defined as the presence of a palpable pulse lasting ≥20 minutes after CPR. The primary outcome measure for this study was comparison of the rate of sustained ROSC (hereafter referred to simply as ROSC).

### Measurements

We collected data on potential covariates, confounders, and effect modifiers relating to patient-specific and CPR-specific factors. Specifically, the patient related data included demographics, severity of critical illness using the Acute Physiology and Chronic Health Evaluation (APACHE) II scoring system,^35^ and chronic disease burden using the Charlson comorbidity index.^36^ The CPR-specific data included initial rhythm, duration of CPR, and use of a mechanical chest compression device. ETCO_2_ and ventilation rate were measured using the Nonin™ RespSense II™, which algorithmically determines ETCO_2_ from a measured CO_2_ waveform and records ventilation rate. To be included, subjects had to have a minimum of 60 seconds of ETCO_2_ and ventilation rate data. As it was possible that some patients had achieved ROSC up to two minutes prior to the last pulse check, we removed the last 120 seconds of ETCO_2_ and ventilation data to remove any potential data that could have reflected ETCO_2_ and ventilation rates immediately post-ROSC when the heart was beating. Patients were grouped into those who were ventilated on average close to the current AHA guideline (6-12 bpm) and those who were ventilated above 12 bpm. This expanded range for the AHA guideline was utilized for consistency with prior studies.^30^ In addition to the average ventilation rate, the overall proportion of CPR time in which a subject was ventilated >12 bpm was calculated. Additionally, data from arterial blood gases collected as part of routine clinical care during the cardiac arrest (hemoglobin concentration, arterial partial pressure of oxygen [PaO_2_] and arterial partial pressure of CO_2_ [PaCO_2_]) were analyzed when clinically drawn by the resuscitation team.

We also sought to understand the dual relationship between ventilation rate and circulation and pulmonary gas exchange. To do so, we first evaluated the relationship between ventilation rate and circulation as measured by ETCO_2_. ETCO_2_ is a routinely used marker of CPR quality since expiratory CO_2_ tension during cardiac arrest is mostly dependent on pulmonary perfusion volume (a proxy for cardiac output).^16^ Second, we evaluated the relationship between ventilatory rate and pulmonary gas exchange as measured by PaO_2_ and PaCO_2_.

The data that support the findings of this study are available from the corresponding author upon reasonable request.

### Statistical Analysis

Data analysis was performed using R software, version 4.2.2, with the significance threshold set at p < 0.05. Dr. Parnia and Ms. Xu had full access to the data and take responsibility for its integrity and the data analysis. Summary statistics were generated for patient demographics and clinical characteristics stratified by ROSC status. Medians with interquartile ranges (IQR) were reported for continuous variables and frequencies with proportions were reported for categorical variables. For continuous variables with >10% missing datapoints, the total number of observations used were reported and the total count of missing data points for categorical variables were reported. Statistical significance was assessed using Wilcoxon rank-sum tests for continuous variables and Pearson’s Chi-squared test or Fisher’s exact test for categorical variables depending on the sample size and distribution. Logistic regression analysis was conducted to assess the impact of ventilation rates for every 2 breaths/min increase on the likelihood of ROSC, adjusting for potential confounding factors including age, gender, initial post-arrest cardiac rhythm, pre-event APACHE-II scores and mechanical chest compression device usage.

A cubic B-spline regression was employed to evaluate for a non-linear relationship between ventilation rates and ROSC outcomes with the optimal breakpoint hyperparameter selected by the lowest Akaike Information Criterion (AIC). A dense grid of 10,000 points was utilized to assess the model’s predictive accuracy across ventilation parameters and visualize the B-spline curve with a 95% confidence band. Potential turning points in the ventilation-ROSC relationship curve were identified to estimate an optimal ventilation rate that maximizes the likelihood of ROSC.

Intra-arrest blood gas (PaO2, PaCO2), and ETCO2 levels between patients ventilated 6-12 breaths/min and patients ventilated > 12 breaths/min, were compared to assess the impact of ventilation rate on cardiac and systemic perfusion, as indicated by pulmonary perfusion dependent ETCO2 levels, and on gas exchange, as reflected by arterial blood gas measurements.

## Results

### Study Population and Patient Characteristics

This study analyzed data from 222 subjects who received capnography monitoring for ventilation rate and ETCO_2_ levels, derived from a total study population of 567 (Table S1). The majority of subjects enrolled in the parent study did not have ETCO_2_ monitoring as this study component was phased into the parent study and was not a required component for all sites. Since subjects only received ventilatory monitoring after advanced airway placement, subjects who had ventilation rate and ETCO_2_ monitoring on average had a longer duration of CPR than those who did not (22 minutes vs. 18 minutes, p = 0.014), but otherwise did not meaningfully differ in terms of demographics, comorbidities, etiology of cardiac arrest, or essential cardiac arrest treatments (Table S1).

Of the 222 included subjects, 89 (40%) achieved sustained ROSC. 11 (4.9%) subjects survived to hospital discharge with good neurologic outcome (Glasgow Outcome Score 4-5), of 14 (6.3%) who survived to hospital discharge.

### ROSC and No ROSC Cohorts

Detailed subject characteristics stratified by ROSC are presented in Table 1. Subjects who achieved ROSC had a higher mean ventilation rate (17.4 breaths/min) than those who did not (15.0 breaths/min; p = 0.002) and spent a higher proportion of their monitored resuscitation time being ventilated greater than 12 breaths/min (80% of the time in those who achieved ROSC vs. 59% in those who did not; p < 0.001). Subjects who achieved ROSC also demonstrated a higher distribution in ETCO_2_ levels, with an 8-mmHg higher median difference in mean ETCO_2_ (25 mm Hg vs. 17 mm Hg, p < 0.001), compared to those who did not achieve ROSC. The ROSC and no ROSC cohorts otherwise did not differ in terms of demographics, comorbidities, etiology of cardiac arrest, or CPR-related interventions (Table 1).

**Table 1.**
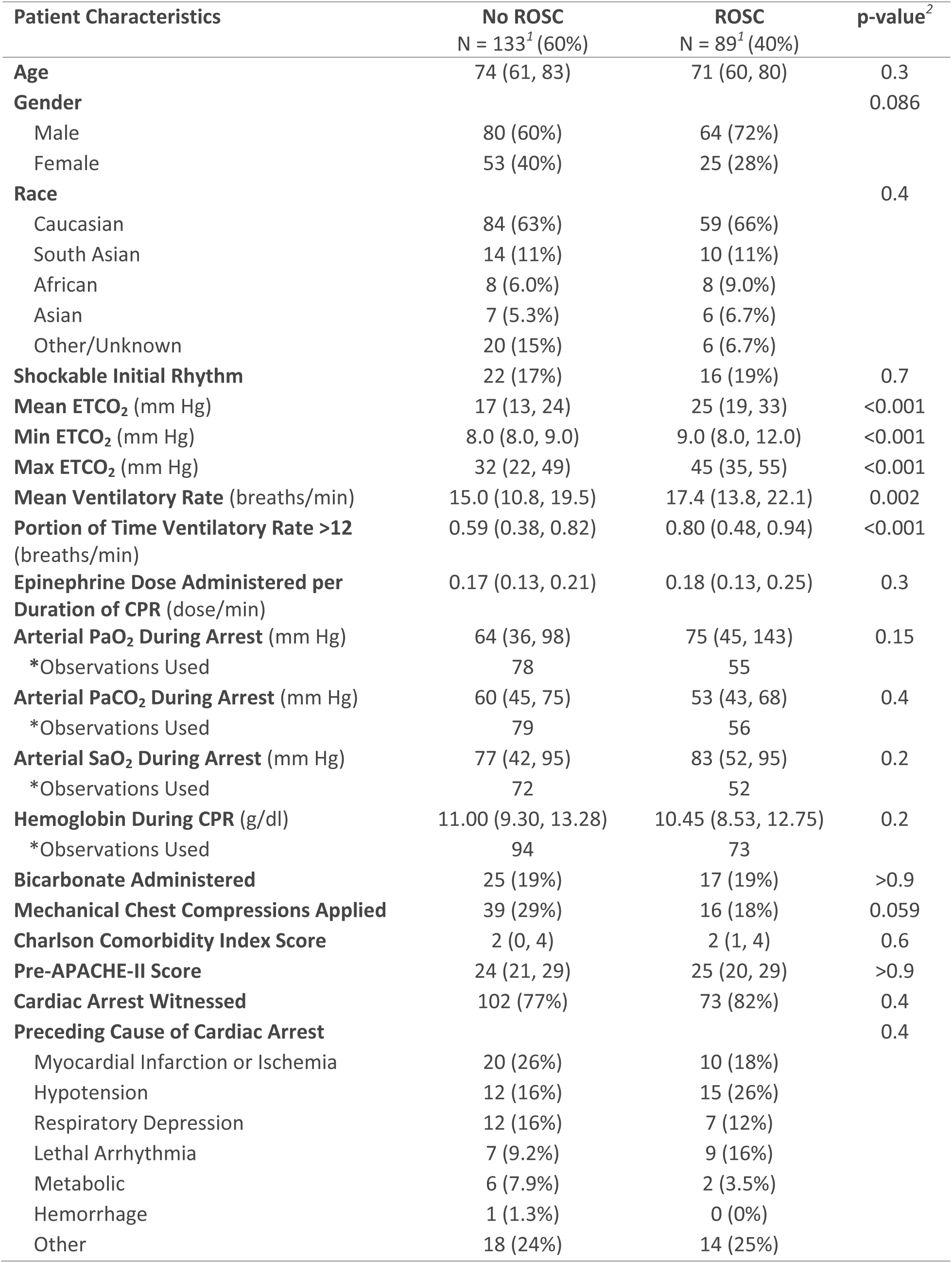
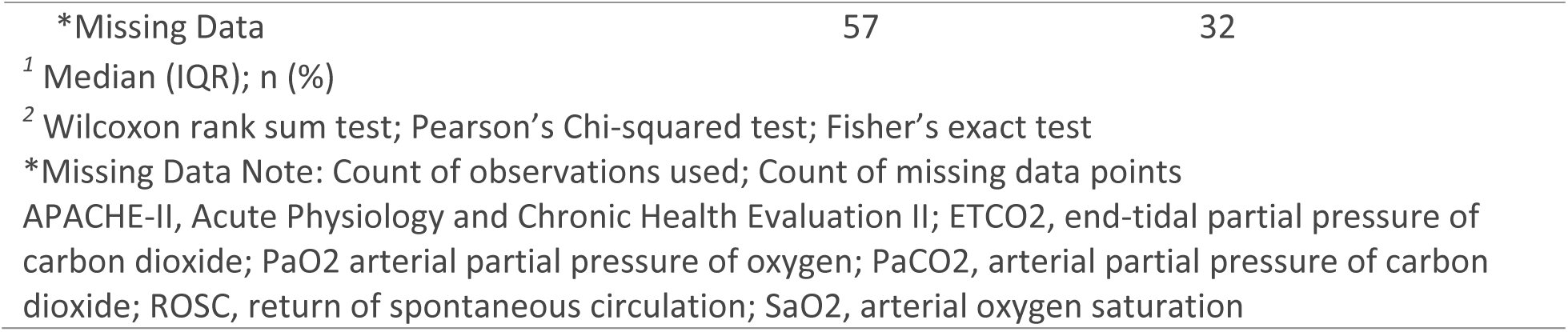
Patient Characteristics Stratified by Return of Spontaneous Circulation (ROSC)

| **Patient Characteristics** | **No ROSC**<br>N = 133<sup>1</sup> (60%) | **ROSC**<br>N = 89<sup>1</sup> (40%) | **p-value<sup>2</sup>** |
| --- | --- | --- | --- |
| **Age** | 74 (61, 83) | 71 (60, 80) | 0.3 |
| **Gender** |  |  | 0.086 |
| Male | 80 (60%) | 64 (72%) |  |
| Female | 53 (40%) | 25 (28%) |  |
| **Race** |  |  | 0.4 |
| Caucasian | 84 (63%) | 59 (66%) |  |
| South Asian | 14 (11%) | 10 (11%) |  |
| African | 8 (6.0%) | 8 (9.0%) |  |
| Asian | 7 (5.3%) | 6 (6.7%) |  |
| Other/Unknown | 20 (15%) | 6 (6.7%) |  |
| **Shockable Initial Rhythm** | 22 (17%) | 16 (19%) | 0.7 |
| **Mean ETCO<sub>2</sub> (mm Hg)** | 17 (13, 24) | 25 (19, 33) | <0.001 |
| **Min ETCO<sub>2</sub> (mm Hg)** | 8.0 (8.0, 9.0) | 9.0 (8.0, 12.0) | <0.001 |
| **Max ETCO<sub>2</sub> (mm Hg)** | 32 (22, 49) | 45 (35, 55) | <0.001 |
| **Mean Ventilatory Rate (breaths/min)** | 15.0 (10.8, 19.5) | 17.4 (13.8, 22.1) | 0.002 |
| **Portion of Time Ventilatory Rate >12 (breaths/min)** | 0.59 (0.38, 0.82) | 0.80 (0.48, 0.94) | <0.001 |
| **Epinephrine Dose Administered per Duration of CPR (dose/min)** | 0.17 (0.13, 0.21) | 0.18 (0.13, 0.25) | 0.3 |
| **Arterial PaO<sub>2</sub> During Arrest (mm Hg)** | 64 (36, 98) | 75 (45, 143) | 0.15 |
| *Observations Used | 78 | 55 |  |
| **Arterial PaCO<sub>2</sub> During Arrest (mm Hg)** | 60 (45, 75) | 53 (43, 68) | 0.4 |
| *Observations Used | 79 | 56 |  |
| **Arterial SaO<sub>2</sub> During Arrest (mm Hg)** | 77 (42, 95) | 83 (52, 95) | 0.2 |
| *Observations Used | 72 | 52 |  |
| **Hemoglobin During CPR (g/dl)** | 11.00 (9.30, 13.28) | 10.45 (8.53, 12.75) | 0.2 |
| *Observations Used | 94 | 73 |  |
| **Bicarbonate Administered** | 25 (19%) | 17 (19%) | >0.9 |
| **Mechanical Chest Compressions Applied** | 39 (29%) | 16 (18%) | 0.059 |
| **Charlson Comorbidity Index Score** | 2 (0, 4) | 2 (1, 4) | 0.6 |
| **Pre-APACHE-II Score** | 24 (21, 29) | 25 (20, 29) | >0.9 |
| **Cardiac Arrest Witnessed** | 102 (77%) | 73 (82%) | 0.4 |
| **Preceding Cause of Cardiac Arrest** |  |  | 0.4 |
| Myocardial Infarction or Ischemia | 20 (26%) | 10 (18%) |  |
| Hypotension | 12 (16%) | 15 (26%) |  |
| Respiratory Depression | 12 (16%) | 7 (12%) |  |
| Lethal Arrhythmia | 7 (9.2%) | 9 (16%) |  |
| Metabolic | 6 (7.9%) | 2 (3.5%) |  |
| Hemorrhage | 1 (1.3%) | 0 (0%) |  |
| Other | 18 (24%) | 14 (25%) |  |

|  |  |  |
| --- | --- | --- |
| *Missing Data | 57 | 32 |
| <sup>1</sup> Median (IQR); n (%) |  |  |
| <sup>2</sup> Wilcoxon rank sum test; Pearson's Chi-squared test; Fisher's exact test |  |  |
| *Missing Data Note: Count of observations used; Count of missing data points |  |  |
| APACHE-II, Acute Physiology and Chronic Health Evaluation II; ETCO<sub>2</sub>, end-tidal partial pressure of carbon dioxide; PaO<sub>2</sub> arterial partial pressure of oxygen; PaCO<sub>2</sub>, arterial partial pressure of carbon dioxide; ROSC, return of spontaneous circulation; SaO<sub>2</sub>, arterial oxygen saturation |  |  |

### Relationship Between Ventilation Rate and ROSC Status

Of patients who had mean ventilation rates > 12 breaths/min, 45% achieved ROSC, while only 24% of patients with ventilation rates near the AHA guideline (6-12 breaths/min) achieved ROSC (p=0.009) (Figure 1). Among the 87 patients who achieved ROSC, 86% of patients had mean ventilation rates over 12 breaths/min while only 14% had ventilation rates in the range of 6-12 breaths/min. Notably, 84.7% of all subjects were ventilated > 10 breaths/min on average.

**Figure 1.**
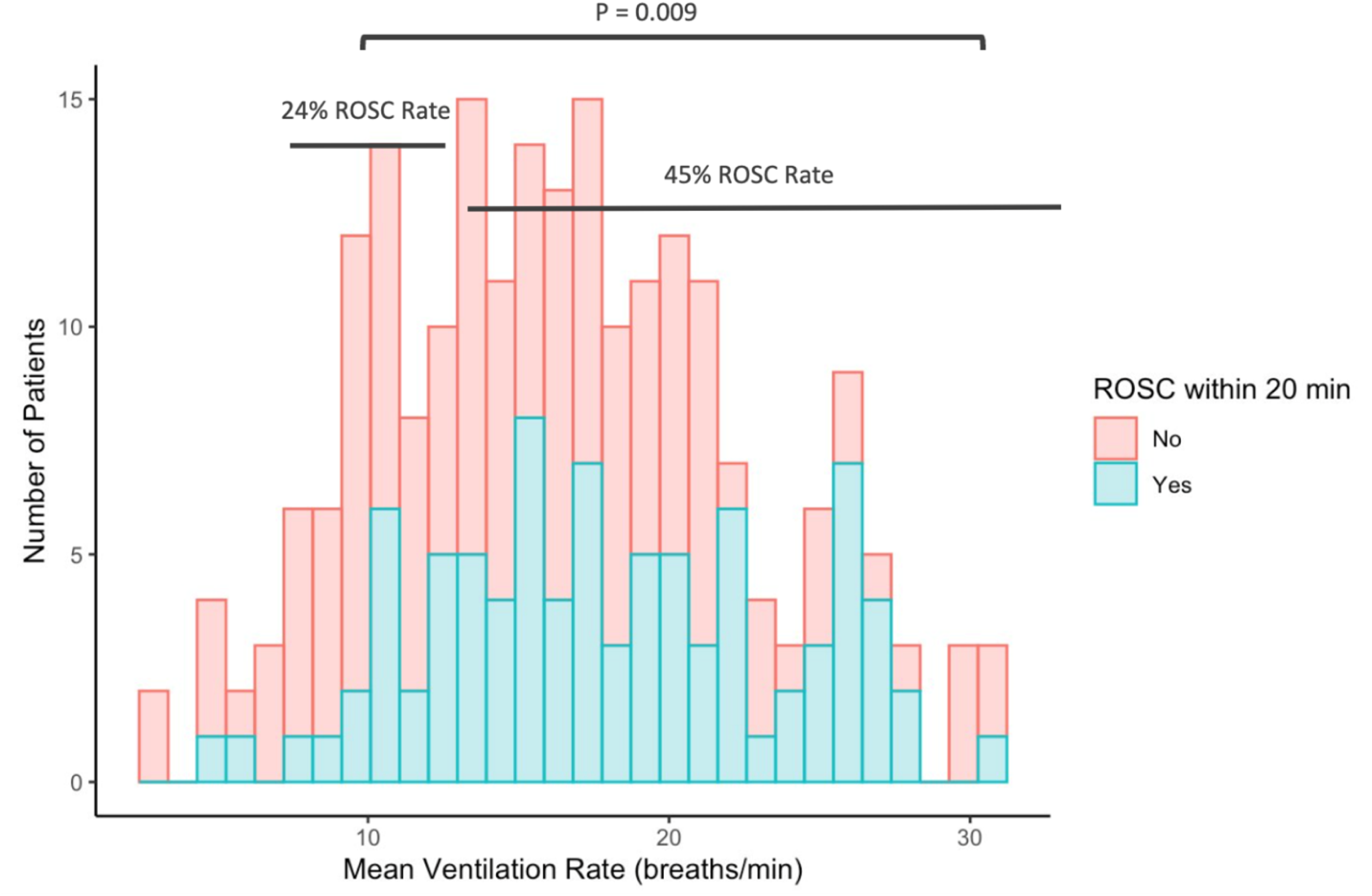
Distribution of Mean Ventilation Rate Stratified by ROSC Outcome. Histogram representing the distribution of mean ventilation rates stratified by patient ROSC outcome status. The proportion of patients who achieved ROSC was computed for subjects ventilated 6-12 breaths/min, and those ventilated > 12 breaths/min. 45% of patients ventilated > 12 breaths/min achieved ROSC, while 24% of patients ventilated 6-12 breaths/min achieved ROSC (p = 0.009).

A two breath/min increase in mean ventilation rate was associated with an increase in the odds of achieving ROSC by a factor of 1.15 (95% CI 1.05 - 1.26, p <0.001 in a univariate model, Table 2). This association was durable in multivariate logistic regression, in which a two breath/min increase in mean ventilation rate was associated with an increase in the odds of achieving ROSC by a factor of 1.15 (95% CI 1.04 - 1.28, p = 0.006), while controlling for age, gender, shockable initial cardiac rhythm, pre-event APACHE II score, and mechanical chest compressions device usage (Table 2).

**Table 2.** Odds Ratio for the Effect of Mean Ventilation Rate on the Likelihood of ROSC.

| Mean Ventilation Rate Model | Odd Ratio (95% CI) | p-value<sup>2</sup> |
| --- | --- | --- |
| Unadjusted | 1.15 (1.05, 1.26) | <0.001 |
| Adjusted for Age, Gender, Shockable Initial Rhythm, Pre-event APACHE II score, and Mechanical Chest Compression Device Usage | 1.15 (1.04, 1.28) | 0.006 |

The adjusted odds ratio is computed for a 2 breaths/min increase in mean ventilation rate. APACHE-II, Acute Physiology and Chronic Health Evaluation II.

A smoothed cubic spline curve demonstrating the relationship between mean ventilation rate and predicted probability of ROSC shows a turning point at the mean ventilation rate of 26.68 breaths/min as a potential threshold of diminishing likelihood of ROSC (Figure 2).

**Figure 2.**
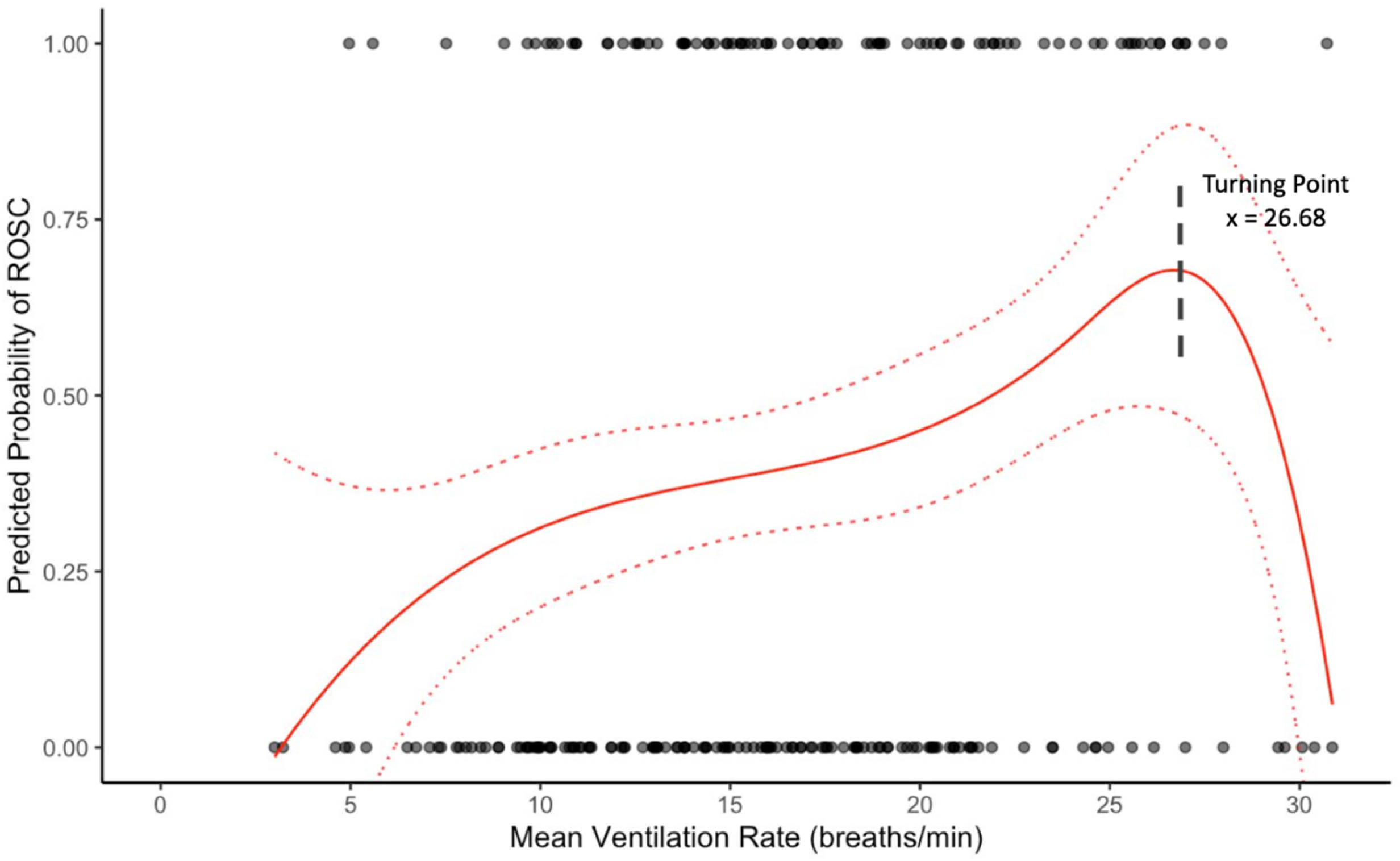
Cubic B-Spline Regression of Mean Ventilation Rate on Predicted Probability of ROSC. Cubic B-spline regression of mean ventilation rate and ROSC with an Akaike Information Criterion (AIC)-optimized internal breakpoint at 24 breaths/min, dense grid of 10,000 points, and 95% CI band. A turning point at 26.68 breaths/min was identified in the ventilation-ROSC relationship curve as a likely optimal mean ventilation rate that maximizes the likelihood of ROSC.

### Impact of Ventilation Rate on Gas Exchange and Perfusion

Patients with mean ventilation rates exceeding 12 breaths/min demonstrated significantly elevated mean ETCO_2_ levels, with a median of 25 mm Hg for patients with mean ventilation rates > 12 breaths/min, marking an 8 mm Hg median increase compared to those with mean ventilation rates within the 6-12 breaths/min range (median = 15 mm Hg; p < 0.001). In order to determine whether this observed difference was potentially driven by differences in minute ventilation due to the discordant ventilation rates, we examined PaO_2_ and PaCO_2_ levels on arterial blood gasses collected during the resuscitation as a part of clinical care. No difference was observed between the ventilation groups for both PaO_2_ and PaCO_2_ levels (Table 3).

**Table 3.** Intra-arrest Blood Gas and ETCO_2_ Levels Distribution Stratified by Mean Ventilation Rate Groups.

| Blood Gas Variable (mm Hg) | Mean Ventilation Rate<br>6-12 breaths/min<br>N = 32 (25%) | Mean Ventilation Rate<br>> 12 breaths/min<br>N = 95<sup>1</sup> (75%) | p-value<sup>2</sup> |
| --- | --- | --- | --- |
| PaCO<sub>2</sub> | 53 (38, 68) | 60 (45, 81) | 0.062 |
| PaO<sub>2</sub> | 75 (37, 90) | 68 (38, 124) | 0.9 |

<sup>1</sup> n (%)
<sup>2</sup> Fisher's exact test; Kruskal-Wallis rank sum test
PaO<sub>2</sub> arterial partial pressure of oxygen; PaCO<sub>2</sub>, arterial partial pressure of carbon dioxide.

## Discussion

We have shown that ventilation rates in cardiac arrest patients with an advanced airway typically exceed AHA guidelines and such cardiac arrest victims who were ventilated at >12 bpm were more likely to achieve ROSC than those ventilated close to the recommended rate (6-12 breaths/min). Notably, our nonlinear regression analysis found this potential benefit may only peak at relatively high ventilation rates (around 26 breaths/min). Additionally, we have shown that in this cohort, higher ventilation rate appeared to improve CPR quality—as demonstrated by ETCO_2_ levels—without adversely impacting pulmonary gas exchange, as indicated by equivalent PaO_2_ and PaCO_2_ levels.

Our findings are consistent with recent guideline-changing evidence that ventilation rates of 20-30 bpm may be beneficial in hospitalized pediatric patients.^28,29^ The authors of that paper hypothesized that children in particular benefit from an elevated ventilation rate since their baseline ventilation rate is higher. Similarly, adult baseline breathing rates are typically cited as 12-20 breaths per minute, which exceeds the current AHA recommendation.^37^ The current AHA guidelines place a considerable emphasis on appropriately prioritizing chest compressions over airway and ventilation, with particular concern placed on the prevention of poor central venous return due to increased intrathoracic pressure from over-ventilation. However, these guidelines were largely based on weak evidence collected from human studies in which compressions and ventilations were alternated, and on animal model studies which did not show 10 breaths/min to be superior to any other studied rate.^27^ Notably, the systematic review (on which the AHA guidelines were largely based) included exclusively studies assessed to be at high risk of bias, and with a total of only 331 human and animal subjects.^13^ Recent evidence from the out-of-hospital setting also supports that hyperventilation in excess of the current guidelines may be beneficial. The Pragmatic Airway Resuscitation Trial (PART) examined 1,010 patients with out-of-hospital cardiac arrest and found that each additional minute of hyperventilation in the 12-16 breaths/min range was associated with increased odds of ROSC (OR: 1.09, 95% CI: 1.04–1.15) and favorable neurological survival (OR: 1.36, 95% CI: 1.01–1.84).^30^

Considering our findings in this context, there would appear to be cause to re-evaluate our current guideline for adult cardiac arrest patients with an advanced airway undergoing continuous compressions and ventilations. A higher ventilation rate range may be superior to the current guideline-recommended rate of 10 breaths/min. Unfortunately, a recent attempt to prospectively test 10 breaths/min vs. 20 breaths/min ventilation rates was halted due to slow recruitment leading to a severely underpowered study,^38^ and to our knowledge, there are no ongoing studies prospectively comparing ventilation rate strategies.

Curiously, we found higher ventilation rates were also associated with increased ETCO_2_. Normally, higher ventilation rates would be expected to increase minute ventilation, which would in turn lead to a decreased ETCO_2_. However, in CPR ETCO_2_ is influenced both by minute ventilation (decreasing with higher minute ventilation) and by the rate at which CO_2_-enriched blood is returned to the lungs (i.e., cardiac output). In this fashion, ETCO_2_ oftentimes serves as a hemodynamic marker in CPR, rather than a ventilatory one. The paradoxically increased ETCO_2_ levels we observed in patients who received greater ventilation rates potentially indicate that higher ventilation rates were associated with increased central venous return and CO_2_ delivery to the lung capillary beds (i.e., cardiac output) compared to lower ventilation rates. Notably, patients who had higher ventilation rates did not have significantly higher PaO_2_ and PaCO_2_. This suggests that effective minute volume may have been similar between these patients and those ventilated at lower rates. A potential explanation for this finding is that patients receiving higher ventilation rates received lower tidal volume respirations, leading to an overall lower intrathoracic pressure during decompression without sacrificing total minute gas exchange. Since elevated intrathoracic pressure is the primary factor implicated in poor venous return to the heart in CPR and resulting hemodynamic compromise, an equivalent minute ventilation delivered over more breaths would understandably lead to increased venous return and CPR-induced cardiac output, which is essential for ROSC.

There is cause to believe that the unique circumstance of continues compressions with continuous ventilations would produce small tidal volumes at higher rates. It is possible that when a manual ventilation attempt occurs simultaneous with a chest compression, the positive intrathoracic pressure generated by the compression would lead to increased ventilation perceived resistance due to the increased peak airway pressure, which may cause the practitioner delivering the ventilation to terminate the ventilation early and resultingly deliver low volume breaths. A recent pilot study of artificial ventilation in prehospital intubated patients demonstrated patients undergoing CPR had a 0.36 ml/kg lower tidal volume (standardized by predicted body weight; 95% CI: 0.13 to 0.58 lower ml/kg) than post-ROSC controls, potentially consistent with this hypothesis.^39^ Unfortunately, a major limitation of our study was that specific ventilation parameters (inspiratory time and tidal volume in particular) were not able to be measured.

A recent study using a porcine model also demonstrated that high-rate, low-volume ventilation (Ultra-Low Tidal Volume Ventilation) provided adequate gas exchange at lower intrathoracic pressures.^40^ It would be logical that during continuous compressions and ventilations in CPR with an advanced airway, healthcare personnel perceive resistance ventilating the patient due to increased intrathoracic pressures during chest compression, naturally leading to higher-rate, lower-volume ventilation. To our knowledge, investigation of a range of intra-arrest ventilation parameters in asynchronous continuous compression and ventilation CPR has not been conducted. Such an investigation certainly appears to be warranted.

Our study has several other limitations. Our ventilation rate was algorithmically computed from waveform capnography, which has previously been shown in other contexts to be potentially susceptible to over-estimation of ventilation rate due to chest compression artifact.^41^ Also, the low neurological survival rate in this cohort (4.9%) and overall survival rate (6.3%) both made survival analysis impossible to adequately undertake. This may be a result of the subjects in this study having a greater severity of illness (with mean pre-arrest APACHE II scores of 23.9, which correlate to an expected hospital mortality rate of >40%).^35^ This may limit the generalizability of our findings to out-of-hospital patients and lower-acuity in-hospital patients. In addition to the subjects being more critically ill at baseline compared to usual populations, the low survival rate observed is likely due to selection bias build into the parent study to enrich for patients in the metabolic phase of cardiac arrest (i.e., the inclusion requirement of >5 minutes of CPR), or because subjects may have received variable-quality CPR. Future studies should also integrate assessment of CPR quality proxies (such as chest compression fraction, chest compression depth, and release velocity), which were not feasible at the time of our original study design. Additionally, while improved odds of ROSC are necessary for improved neurological survival outcomes, they are certainly not sufficient. Future studies adequately powered to examine survival are necessary. Furthermore, studies have suggested that the movement from ROSC to neurological survival is mediated by post-resuscitation care, especially for patients who reached the metabolic phase of cardiac arrest.^42^ Future studies will need to incorporate these post-resuscitation variables as well.

In conclusion, in this study of in-hospital cardiac arrests with an advanced airway, ventilation in excess of AHA guidelines was common. Ventilation rates >12 breaths/min were associated with increased odds of ROSC, though ventilation rates >26 breaths/min were steeply associated with decreased odds of ROSC. Current CPR guidelines may need to be reevaluated, and considerations made for allowing for a higher ventilation rate in intubated patients undergoing continuous asynchronous compressions and ventilation, although further research on this relationship and the impact of ventilation rate on survival is needed. Although higher rates of ROSC may be due to the inadvertent delivery of high-rate, low-volume ventilation, which has some supporting translational evidence, further research explicitly examining ventilation parameters would be required to fully evaluate this theory. Future studies examining ventilation rates with inclusion of CPR quality metrics and specific monitoring of ventilation parameters would be very informative.

## Non-Standard Abbreviations

AWARE II: Awareness During Resuscitation II
CPR: cardiopulmonary resuscitation
ETCO2: end-tidal partial pressure of carbon dioxide
PaCO2: arterial partial pressure of carbon dioxide
PaO2: arterial partial pressure of oxygen
ROSC: return of spontaneous circulation
SaO2: arterial oxygen saturation

## References

1. Tsao CW, Aday AW, Almarzooq ZI, Alonso A, Beaton AZ, Bittencourt MS, Boehme AK, Buxton AE, Carson AP, Commodore-Mensah Y. Heart disease and stroke statistics—2022 update: a report from the American Heart Association. Circulation. 2022;145:e153–e639. doi:

2. Andersen LW, Holmberg MJ, Berg KM, Donnino MW, Granfeldt A. In-hospital cardiac arrest: a review. Jama. 2019;321:1200–1210. doi:

3. Peberdy MA, Ornato JP, Larkin GL, Braithwaite RS, Kashner TM, Carey SM, Meaney PA, Cen L, Nadkarni VM, Praestgaard AH. Survival from in-hospital cardiac arrest during nights and weekends. Jama. 2008;299:785–792. doi:

4. Girotra S, Nallamothu BK, Spertus JA, Li Y, Krumholz HM, Chan PS. Trends in survival after inhospital cardiac arrest. New England Journal of Medicine. 2012;367:1912–1920. doi:

5. Song F, Sun S, Ristagno G, Yu T, Shan Y, Chung SP, Weil MH, Tang W. Delayed high-quality CPR does not improve outcomes. Resuscitation. 2011;82:S52–S55. doi:

6. Wong CX, Brown A, Lau DH, Chugh SS, Albert CM, Kalman JM, Sanders P. Epidemiology of sudden cardiac death: global and regional perspectives. *Heart*, Lung and Circulation. 2019;28:6–14. doi:

7. Riva G, Ringh M, Jonsson M, Svensson L, Herlitz J, Claesson A, Djärv T, Nordberg P, Forsberg S, Rubertsson S. Survival in out-of-hospital cardiac arrest after standard cardiopulmonary resuscitation or chest compressions only before arrival of emergency medical services: nationwide study during three guideline periods. Circulation. 2019;139:2600–2609. doi:

8. Ofoma UR, Basnet S, Berger A, Kirchner HL, Girotra S. Trends in survival after in-hospital cardiac arrest during nights and weekends. Journal of the American College of Cardiology. 2018;71:402–411. doi:

9. Svensson L, Bohm K, Castrèn M, Pettersson H, Engerström L, Herlitz J, Rosenqvist M. Compression-only CPR or standard CPR in out-of-hospital cardiac arrest. New England Journal of Medicine. 2010;363:434–442. doi:

10. Rössler B, Goschin J, Maleczek M, Piringer F, Thell R, Mittlböck M, Schebesta K. Providing the best chest compression quality: Standard CPR versus chest compressions only in a bystander resuscitation model. PloS one. 2020;15:e0228702. doi:

11. Wang PL, Brooks SC. Mechanical versus manual chest compressions for cardiac arrest. Cochrane Database of Systematic Reviews. 2018. doi:

12. Zhan L, Yang LJ, Huang Y, He Q, Liu GJ. Continuous chest compression versus interrupted chest compression for cardiopulmonary resuscitation of non-asphyxial out-of-hospital cardiac arrest. Cochrane Database of Systematic Reviews. 2017. doi:

13. Vissers G, Soar J, Monsieurs KGJR. Ventilation rate in adults with a tracheal tube during cardiopulmonary resuscitation: a systematic review. 2017;119:5–12. doi:

14. Orso D, Vetrugno L, Federici N, Borselli M, Spadaro S, Cammarota G, Bove T. Mechanical ventilation management during mechanical chest compressions. Respiratory Care. 2021;66:334–346. doi:

15. Cordioli RL, Grieco DL, Charbonney E, Richard J-C, Savary D. New physiological insights in ventilation during cardiopulmonary resuscitation. Current opinion in critical care. 2019;25:37–44. doi:

16. Panchal AR, Bartos JA, Cabañas JG, Donnino MW, Drennan IR, Hirsch KG, Kudenchuk PJ, Kurz MC, Lavonas EJ, Morley PT. Part 3: Adult Basic and Advanced Life Support: 2020 American Heart Association Guidelines for Cardiopulmonary Resuscitation and Emergency Cardiovascular Care. Circulation. 2020;142:S366–S468. doi:

17. Neth MR, Benoit JL, Stolz U, McMullan J. Ventilation in Simulated Out-of-Hospital Cardiac Arrest Resuscitation Rarely Meets Guidelines. Prehospital Emergency Care. 2020:1–9. doi:

18. Losert H, Sterz F, Köhler K, Sodeck G, Fleischhackl R, Eisenburger P, Kliegel A, Herkner H, Myklebust H, Nysæther J. Quality of cardiopulmonary resuscitation among highly trained staff in an emergency department setting. Archives of internal medicine. 2006;166:2375–2380. doi:

19. O’Neill JF, Deakin CD. Do we hyperventilate cardiac arrest patients? Resuscitation. 2007;73:82–85. doi:

20. Maertens VL, De Smedt LE, Lemoyne S, Huybrechts SA, Wouters K, Kalmar AF, Monsieurs KG. Patients with cardiac arrest are ventilated two times faster than guidelines recommend: an observational prehospital study using tracheal pressure measurement. Resuscitation. 2013;84:921–926. doi:

21. Aufderheide TP, Lurie KG. Death by hyperventilation: a common and life-threatening problem during cardiopulmonary resuscitation. Critical care medicine. 2004;32:S345–S351. doi:

22. Aufderheide TP, Sigurdsson G, Pirrallo RG, Yannopoulos D, McKnite S, Von Briesen C, Sparks CW, Conrad CJ, Provo TA, Lurie KGJC. Hyperventilation-induced hypotension during cardiopulmonary resuscitation. 2004;109:1960–1965. doi:

23. Yannopoulos D, McKnite SH, Tang W, Zook M, Roussos C, Aufderheide TP, Idris AH, Lurie KG. Reducing ventilation frequency during cardiopulmonary resuscitation in a porcine model of cardiac arrest. Respiratory care. 2005;50:628–635. doi:

24. Pirrallo RG, Aufderheide TP, Provo TA, Lurie KG. Effect of an inspiratory impedance threshold device on hemodynamics during conventional manual cardiopulmonary resuscitation. Resuscitation. 2005;66:13–20. doi:

25. Idris AH, Becker LB, Fuerst RS, Wenzel V, Rush WJ, Melker RJ, Orban DJ. Effect of ventilation on resuscitation in an animal model of cardiac arrest. Circulation. 1994;90:3063–3069. doi:

26. Neth MR, Idris A, McMullan J, Benoit JL, Daya MRJJotACoEPO. A review of ventilation in adult out-of-hospital cardiac arrest. 2020;1:190–201. doi:

27. Vissers G, Duchatelet C, Huybrechts S, Wouters K, Hachimi-Idrissi S, Monsieurs KJR. The effect of ventilation rate on outcome in adults receiving cardiopulmonary resuscitation. 2019;138:243–249. doi:

28. Sutton RM, Reeder RW, Landis WP, Meert KL, Yates AR, Morgan RW, Berger JT, Newth CJ, Carcillo JA, McQuillen PS. Ventilation rates and pediatric in-hospital cardiac arrest survival outcomes. Critical care medicine. 2019;47:1627–1636. doi:

29. Topjian AA, Raymond TT, Atkins D, Chan M, Duff JP, Joyner Jr BL, Lasa JJ, Lavonas EJ, Levy A, Mahgoub M. Part 4: pediatric basic and advanced life support: 2020 American Heart Association guidelines for cardiopulmonary resuscitation and emergency cardiovascular care. Circulation. 2020;142:S469–S523. doi:

30. Wang HE, Jaureguibeitia X, Aramendi E, Nichol G, Aufderheide T, Daya MR, Hansen M, Nassal M, Panchal AR, Nikolla DA. Airway strategy and ventilation rates in the pragmatic airway resuscitation trial. Resuscitation. 2022;176:80–87. doi:

31. Parnia S, Shirazi TK, Patel J, Tran L, Sinha N, O’Neill C, Roellke E, Mengotto A, Findlay S, McBrine M. AWAreness during REsuscitation-II: A multi-center study of consciousness and awareness in cardiac arrest. Resuscitation. 2023;191:109903. doi:

32. Weisfeldt ML. A three phase temporal model for cardiopulmonary resuscitation following cardiac arrest. Transactions of the American Clinical and Climatological Association. 2004;115:115. doi:

33. Neumar RW, Otto CW, Link MS, Kronick SL, Shuster M, Callaway CW, Kudenchuk PJ, Ornato JP, McNally B, Silvers SM. Part 8: adult advanced cardiovascular life support: 2010 American Heart Association guidelines for cardiopulmonary resuscitation and emergency cardiovascular care. Circulation. 2010;122:S729–S767. doi:

34. Link MS, Berkow LC, Kudenchuk PJ, Halperin HR, Hess EP, Moitra VK, Neumar RW, O’Neil BJ, Paxton JH, Silvers SM. Part 7: adult advanced cardiovascular life support: 2015 American Heart Association guidelines update for cardiopulmonary resuscitation and emergency cardiovascular care. Circulation. 2015;132:S444–S464. doi:

35. Knaus WA, Draper EA, Wagner DP, Zimmerman JE. APACHE II: a severity of disease classification system. Critical care medicine. 1985;13:818–829. doi:

36. Charlson M, Szatrowski TP, Peterson J, Gold J. Validation of a combined comorbidity index. Journal of clinical epidemiology. 1994;47:1245–1251. doi:

37. Yuan G, Drost NA, McIvor RA. Respiratory rate and breathing pattern. McMaster Univ Med J. 2013;10:23–25. doi:

38. Prause G, Zoidl P, Eichinger M, Eichlseder M, Orlob S, Ruhdorfer F, Honnef G, Metnitz PG, Zajic P. Mechanical ventilation with ten versus twenty breaths per minute during cardio-pulmonary resuscitation for out-of-hospital cardiac arrest: A randomised controlled trial. Resuscitation. 2023;187:109765. doi:

39. Yang BY, Blackwood JE, Shin J, Guan S, Gao M, Jorgenson DB, Boehl JE, Sayre MR, Kudenchuk PJ, Rea TD. A pilot evaluation of respiratory mechanics during prehospital manual ventilation. Resuscitation. 2022;177:55–62. doi:

40. Ruemmler R, Ziebart A, Moellmann C, Garcia-Bardon A, Kamuf J, Kuropka F, Duenges B, Hartmann EK. Ultra-low tidal volume ventilation—A novel and effective ventilation strategy during experimental cardiopulmonary resuscitation. Resuscitation. 2018;132:56–62. doi:

41. Leturiondo M, de Gauna SR, Ruiz JM, Gutiérrez JJ, Leturiondo LA, González-Otero DM, Russell JK, Zive D, Daya M. Influence of chest compression artefact on capnogram-based ventilation detection during out-of-hospital cardiopulmonary resuscitation. Resuscitation. 2018;124:63–68. doi:

42. Granfeldt A, Andersen LW. The new era of post-resuscitation care. Resuscitation. 2022;171:98–99. doi:

